# Investigating the effect of biomass fuel use and Kitchen location on Maternal Report of Birth size: A Cross-Sectional Analysis of 2016 Ethiopian Demographic Health Survey data

**DOI:** 10.1101/2020.09.19.20197871

**Authors:** Girum Gebremeskel Kanno, Adane Tesfaye Anbesse, Mohammed Feyisso Shaka, Miheret Tesfu Legesse, Sewitemariam Desalegn Andarge

## Abstract

Household air pollution from the use of biomass fuels has been associated with low birth weight in many developing countries. We investigated the effect of indoor air pollution from biomass fuel use and kitchen location on maternal reports of birth size in newborn children in Ethiopia using secondary cross-sectional data from Ethiopian Demographic Health Survey conducted in 2016. Birth weight from child health card and/or mothers’ recall was the dependent dichotomous variable. Fuel type was classified as high pollution fuels, and low pollution fuels. Hierarchical logistic regression was used to assess the effect of fuel type on birth weight. The prevalence of low birth weight was 25.9% and the use of biomass fuels was significantly associated with higher odds of having low birth weight baby in the bivariate analysis, after controlling for child and maternal factors. In the final model, the association turned insignificant with AOR, 1.3 (95% 0.9, 1.9). The kitchen location, Gender of the baby, Mother’s Anemia Status, Maternal Chat chewing, and wealth Index were significant factors in the final model. The use of biomass fuels and kitchen location were associated with reduced child size at birth. Further observational studies should investigate this association using more direct methods.

**Practical Implications:** The findings from this study have important implication at national level for policy makers. Ethiopia is a country with a huge proportion of the population depends on biomass fuels for cooking. Despite the progress made to reduce the burden of maternal and infant mortality and morbidity in the pre SDG era, LBW is still one of the challenges that need to be addressed. Identifying the link between biomass fuel use and kitchen location with low birth weight can help the efforts to revise, amend or implement programs that help achieve the SDG goal six, by engaging the energy and health sectors together.

## Introduction

Low birth weight is defined as weight at birth less than 2500 grams. It is a significant public health problem globally and is associated with a range of both short and long term consequences [1]. It is an important marker of maternal and fetal health and nutritional status [2]. Worldwide 15% to 20% of all births are low birth weight, representing more than 20 million births a year [3]. The great majority of low birth weight births occur in low- and middle-income countries especially in the most vulnerable populations, with regional estimates of 13% in sub-Saharan Africa and 26% in Ethiopia [1, 4]. In most developing countries including Ethiopia, the data on low birth weight remain limited or unreliable, as many births occur in homes or small health facilities and mostly they are underreported or not reported at all in official figures, where the final consequence is underestimation of the prevalence of low birth weight [5].

Low birth weight is a major predictor of prenatal mortality and morbidity and compared with babies born at or above the low birth weight cut-off (2,500 g), babies born with low birth weight have a higher risk of stunting, lower IQ, and a higher chance of death during their childhood and also have increased risk for non-communicable diseases such as diabetes and cardiovascular disease later in life [6-9].

Safe environment is one of the basic needs for mothers to grow a healthy baby along with good nutrition, rest, and adequate antenatal care [10]. Different studies have indicated that low birth weight has been a result of exposure to unsafe indoor environment which is mainly the result of household air pollution resulting from cooking fuels and environmental tobacco smoke (ETS) in most developing countries [11-12]. Combustion from these solid fuels in simple household cookstoves contributes to household air pollution (HAP) by emitting considerably large amounts of noxious pollutants and health-damaging airborne pollutants, including particulate matter (PM), carbon monoxide (CO), nitrogen dioxide (NO_2_) formaldehyde, and many other toxic polycyclic aromatic hydrocarbons (PAHs) [13-16]. Out of this noxious pollutants, Carbon monoxide is a well-known feto-toxicant chemical associated with poor fetal growth. The mechanism could be in two ways. The first mechanism occurs when the amount of oxygen supply that must be delivered to tissues have decreased and phenomenon called hypoxia occurred because carbon monoxide interacts with hemoglobin to cross the placenta, which limits the ability of the placenta to transfer nutrients to the fetus. The other occurs when inhaled particulate matter from smoke impairs fetal growth by damaging cells through oxidative stress. [17-20].

Almost 3 billion people, mainly in low and middle-income countries, and 90% of the rural household population in developing countries still rely on high polluting solid fuels (wood, animal dung, charcoal, crop wastes, and coal) burnt in inefficient, highly polluting stoves for cooking and heating and lighting, which are responsible for producing a high concentration of particulate matter in the indoor environment [15-16, 22].

In sub-Saharan Africa the number of people with biomass fuel use has showed no significant change in three decades time from 1980 to 2010, yet the population exposed to indoor air pollution has increased from 333 million to 646 million [23]. This can be translated as, 76% of particulate matter air pollution around the world occurs indoors in developing countries. When biomass fuels are burnt on traditional, typically simple, inefficient, and unwanted household cooking stoves, they producing large volumes of indoor smoke or air pollutant which exceed the safe levels recommended by the World Health Organization which is (recommended 24-hour mean: PM_2.5_ < 25 μg/m^3^ and PM_10_ <50 μg/m^3^) [15, 16, 23-24].

In the sub-Saharan Africa region, where Ethiopia is part of, the leading risk factor for neonatal death (which accounts for more than half of under-five mortality) is low birth weight [25]. The global under-five mortality data indicated that in order to achieve the sustainable development goal (SDG) target of 25 per 1000 live births by 2030 in sub-Saharan Africa (SSA) region, interventions must be enhanced to change the current situations [26].

In Ethiopia, more than 95% of households rely on biomass fuels for cooking, and in almost 53 % of households, food is cooked inside the house creating a favorable condition for indoor air pollution, the largest single environmental risk factor for adverse pregnancy outcomes such as premature death and low birth weight [4, 22].

Different studies conducted in developing countries such as India, Bangladesh, Pakistan, and Malawi, indicated that the use of high polluting cooking fuels was associated with LBW [12, 27-30]. Most of the previously conducted studies in Ethiopia that assessed the predictors of low birth weight mainly focused on maternal, child, and socio-demographic related factors only [31-34], where maternal exposure to indoor air pollution was not taken into consideration while other studies such as [35-36] tried to assess the impact of different fuel types on child size at birth but their focus was localized to the small study area. Another study also assessed the predictors of being small size at Birth using the 2011 EDHS the nation-wide data [37], but the impact of the type of household fuel types and the kitchen location on the birth weight of the child at birth was not included. A starting point to fill the gap is the provision of national empirical evidence on the magnitude of the risk posed by IAP to LBW in the Ethiopian context. Therefore, this study aimed to assess the relationship between exposure to maternal biomass fuel use and low birth weight at the national level in Ethiopia. In this study, in addition to these factors, we tried to determine the effect of biomass fuel use and kitchen location on birth size among births in the last five years proceeding to 2016 EDHS.

## Materials and methods

### Data Source, Setting, and Study Design

Data for this cross-sectional study was obtained from secondary data of 2016 EDHS. The census frame is a complete list of 84,915 enumeration areas (EAs) created for the 2007 PHC. Two-stage stratified sampling was applied to identify eligible residential households across 645 enumeration areas (EAs). Each region was stratified into urban and rural areas, yielding 21 sampling strata. Samples of EAs were selected independently in each stratum in two stages. Implicit stratification and proportional allocation were achieved at each of the lower administrative levels by sorting the sampling frame within each sampling stratum before sample selection, according to administrative units at different levels, and by using a probability proportional to size selection at the first stage of sampling. In the first stage, a total of 645 EAs (202 in urban areas and 443 in rural areas) were selected with probability proportional to EA size (based on the 2007 PHC) and with independent selection in each sampling stratum. The resulting lists of households served as a sampling frame for the selection of households in the second stage. In the EDHS 2016 survey, the total number of births reported in the previous five years was 44,596.

### Dependent Variable/Outcome variable

Our dependent variable was the maternal reported birth size. In Ethiopia as in most developing countries, the majority of deliveries take place at home, and as a result information on birth weight was obtained only for small (14% of births) of babies which were weighed at birth [4]. Information on birth weight was collected by either a written record or the mother’s recall on the size of their babies. The mother’s assessment of the child’s weight was critical because information on birth weight was rarely available for most of the births in the last five years. Respondents (mothers) were asked, “At birth, what was the size of the baby?” The options were ‘very large’, ‘larger than average’, ‘average’, ‘smaller than average’, and ‘very small’. An infant was classified as being LBW (<2500 gram) if the mother reported that they were ‘very small’ or ‘smaller than average’. The rest were labeled as not low birth weight (>2500 gram). Births with missing information about the size of the baby at birth (75.3%) and multiple births (8.2%) were excluded from our analysis.

### Exposure variables

The main independent variables of interest for this study were cooking fuel type and Kitchen location. The standard DHS used eleven fold classification of cooking fuels used in the house. The specific questions asked were, “What type of fuel does your household mainly use?” “Is the cooking usually done in the house, in a separate building, or outdoors?” For our analysis, the main cooking fuel used was grouped into two namely; high pollution cooking fuels (wood, straw, animal dung, and crop residues, kerosene, coal, and charcoal), and low pollution cooking fuels (electricity, liquid petroleum gas, natural gas, and biogas). On the other hand kitchen location was categorized into three groups; food cooked “in the house”, “in a separate building” and “outdoors”.

### Other predictor variables

Other independent variables included in the model were maternal age in years (<20, 20-29, and 30-49 years), maternal education (none, primary, secondary, higher), maternal body mass index (BMI) (Underweight, Normal, Overweight or Obese), wealth index (1 to 5 from poorest to richest, calculated based on the availability of household assets using principal component analysis and provided in the dataset) [4], Birth order (first, second, third, fourth or higher), Gender of the child (Male, Female), Pregnancy intention (planned, mistimed, unplanned), Residence (urban, rural), Chat Chewing (No, Yes) and Alcohol drinking (No, Yes).

We categorized a BMI of less than 18.5 as underweight, 18.5 to less than 25.0 as normal, 25.0 to 29.0 as overweight, and greater than 29.0 as Obese [4]. We used the wealth index quintiles reported in the EDHS data, which is constructed using principal components analysis on the possession of different household assets, adjusting for urban-rural differences [4]. Pregnancy intention was categorized into three groups based on whether the birth or pregnancy was wanted then (planned), wanted later (mistimed), or unwanted (unplanned) at the time of conception.

### Inclusion and exclusion criteria

A total of 44,596 births were reported during the previous five years and birth size data was available only for 11,023 of the births either from card or mother’s recall. Out of these 10,730 cases were singleton births and 10,014 cases have fulfilled the inclusion criteria as indicated in (Fig 1). Apart from birth size information and twin cases, those cases with missing values from child-related, maternal, and household factors were excluded. Cigarette smoking mothers were excluded from this study because they were very low in number and all the smokers were found to be in the ‘high polluting cooking fuel users’ category.

**Fig 1.**
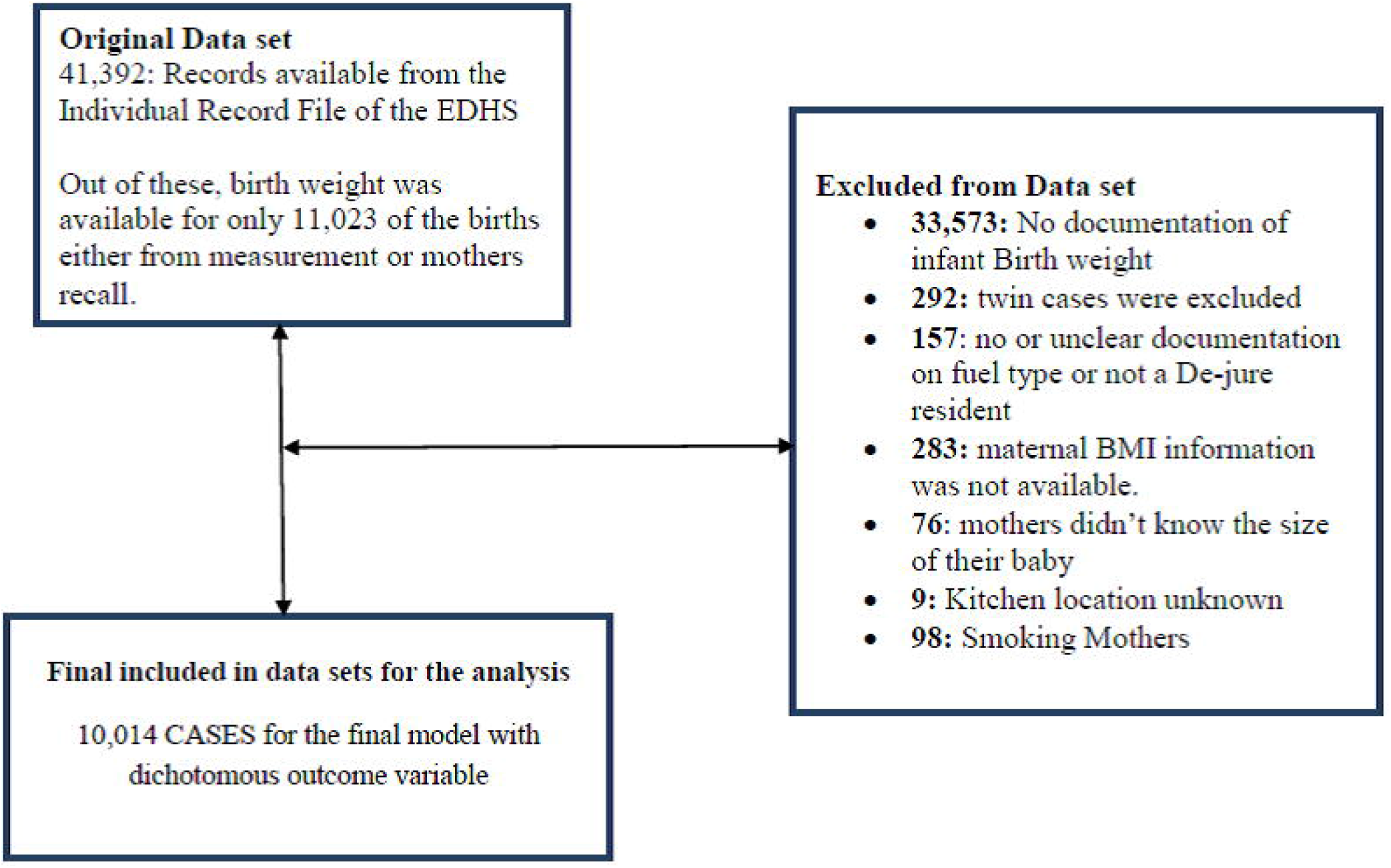
Included and excluded cases on data management process.

### Data analysis

For the bivariate analysis of categorical variables, we used binary logistic regression. We carried out the analyses using SPSS Version 20 software. In addition to the type of fuel used, other (independent) variables considered were child factors (gender of the baby, and birth order), maternal factors (Anemia level, BMI, age at first childbirth, Chat chewing, Alcohol drinking, education, and pregnancy intention) and socio-demographic factors (place of residence (urban/rural) wealth index, sex of head of the household). The collinearity test was done to check the possibility of the presence of collinearity using Pearson’s correlation matrix and Pearson’s correlation coefficients were calculated to rule out multicollinearity. Multilevel modeling was done to adjust for cluster sampling (cluster as the primary sampling unit used in DHS). During statistical modeling, certain factors known to confound the results were adjusted. Hosmer and Lemeshow test was used to check for model fitness. Adjusted odds ratios (AOR) and their 95% confidence intervals (95% CI) were calculated. A p-value of less than 0.05 was considered significant.

### Ethics Statement

This secondary analysis was exempted from ethical review approval, because it used publicly available, de-identified data. However, a request to access datasets from DHS was made, and a letter of permission to use the data set was obtained before the analysis was made.

## Results

### Descriptive statistics

A total of 44,596 births were reported during the previous five years and birth size data was available only for 11,023, of the births and only 10,014 cases were considered for this analysis. According to the mother’s report of the child’s size at birth, 1602(16.0%) were very small, 989(9.9%) smaller than average and 74.1% were average or larger, which is more or less similar with the overall EDHS 2016 report, which shows 16% of births were very small, 10% were smaller than average, and 73% were average or larger. The proportion of infants which were LBW was 2591 (25.9%). Of these 2591 (25.9%) LBW infants, the majority 2538(97.9%) were from households with biomass fuel users (such as charcoal, wood, straws or crops ad animal dungs), and in 1181(45.6%) of the households food has been cooked inside the house. The dominant types of fuel used by the households with newborns in Ethiopia was wood (8383 83.7%) and only about three percent (274) of households use electricity [Table 1].

**Table 1.**
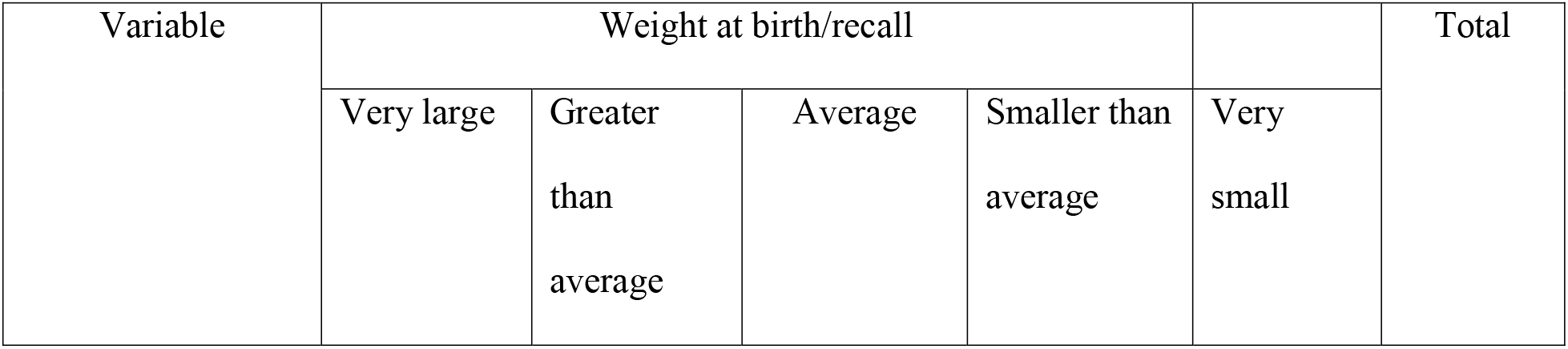
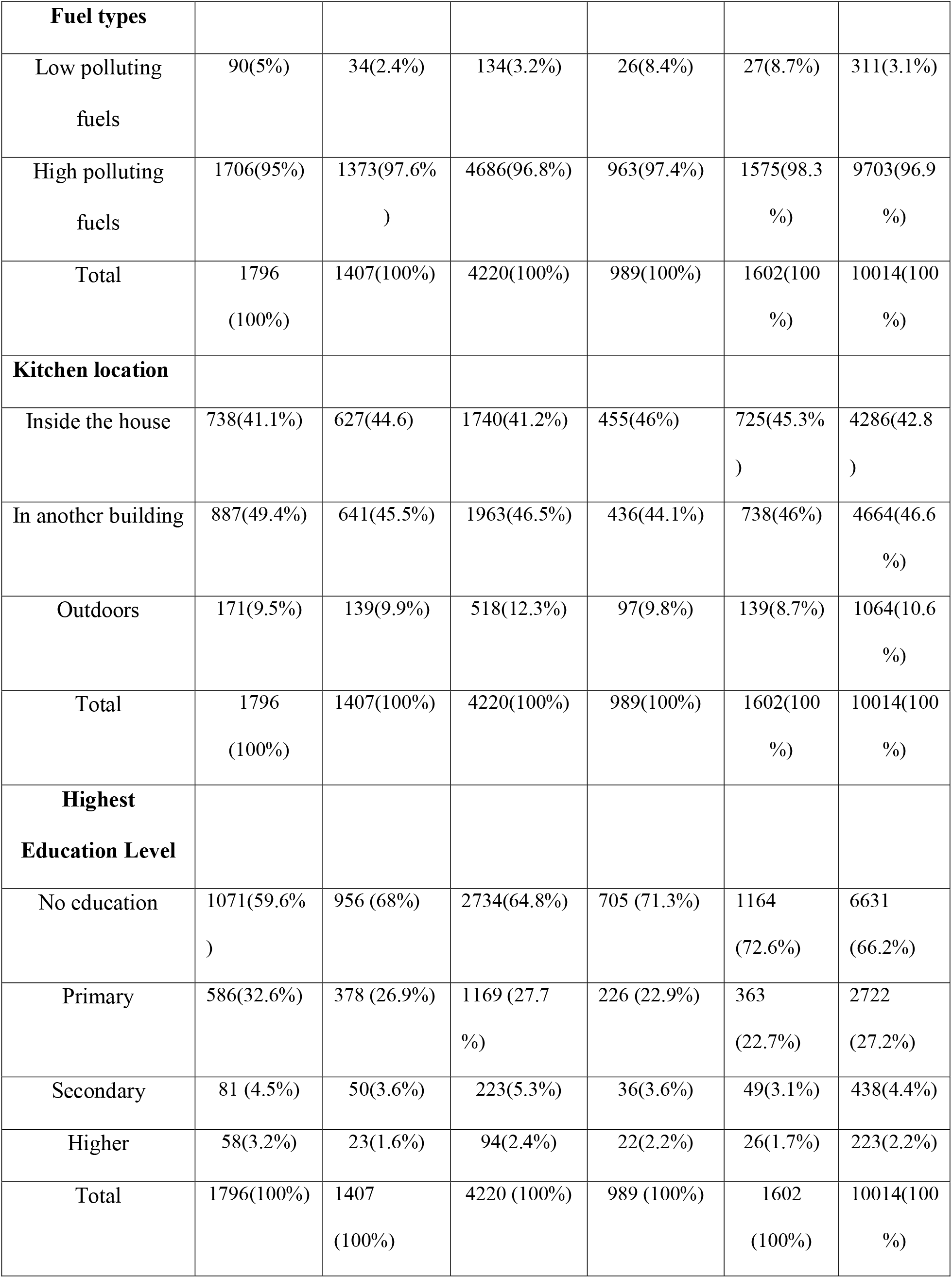
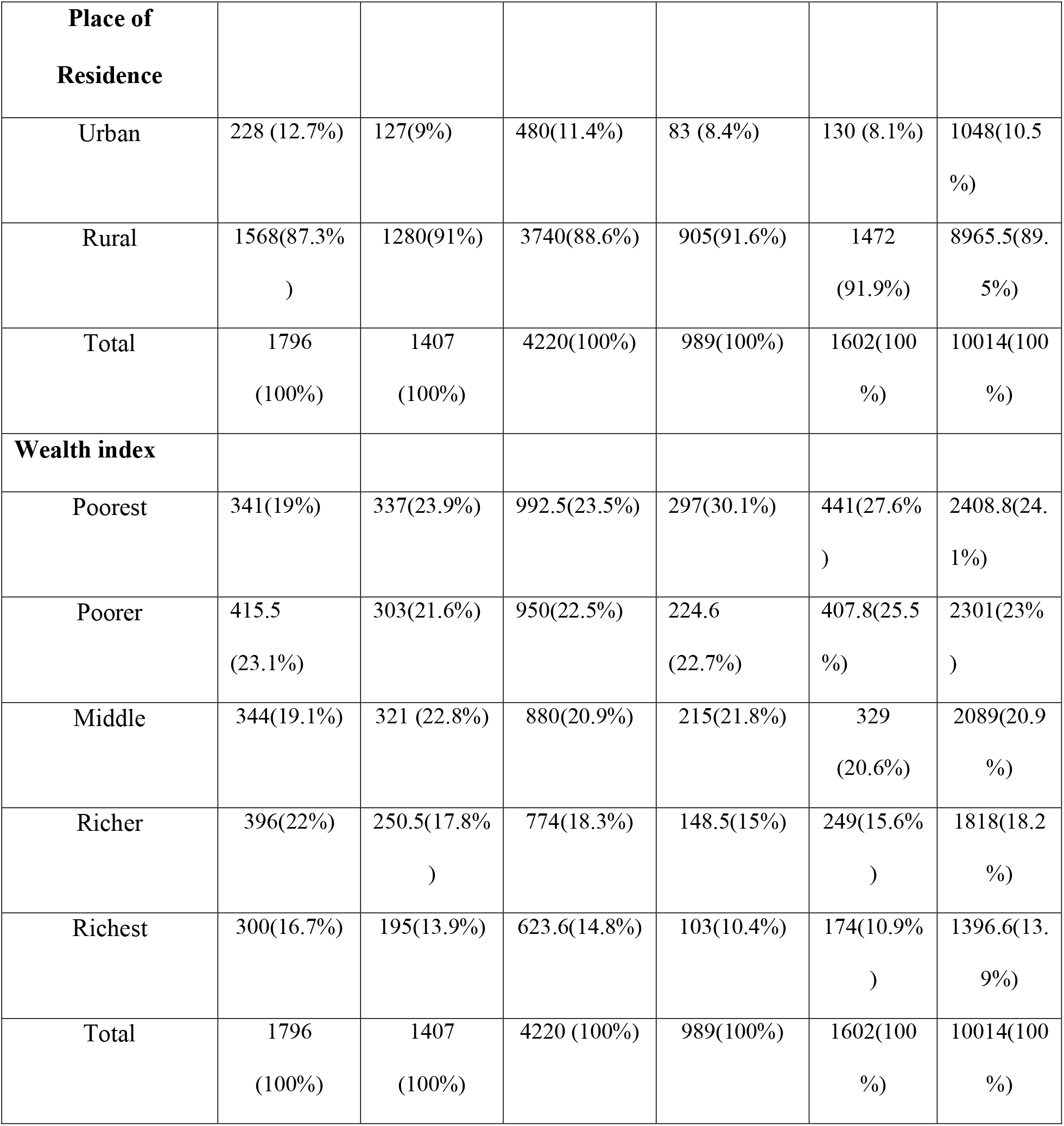
Distribution of birth sizes according to biomass fuel use, Kitchen location and selected socio-demographic factors.

| Variable | Weight at birth/recall |  |  |  |  | Total |
| --- | --- | --- | --- | --- | --- | --- |
|  | Very large | Greater than average | Average | Smaller than average | Very small |  |
| **Fuel types** |  |  |  |  |  |  |
| Low polluting fuels | 90(5%) | 34(2.4%) | 134(3.2%) | 26(8.4%) | 27(8.7%) | 311(3.1%) |
| High polluting fuels | 1706(95%) | 1373(97.6%) | 4686(96.8%) | 963(97.4%) | 1575(98.3%) | 9703(96.9%) |
| Total | 1796(100%) | 1407(100%) | 4220(100%) | 989(100%) | 1602(100%) | 10014(100%) |
| **Kitchen location** |  |  |  |  |  |  |
| Inside the house | 738(41.1%) | 627(44.6%) | 1740(41.2%) | 455(46%) | 725(45.3%) | 4286(42.8%) |
| In another building | 887(49.4%) | 641(45.5%) | 1963(46.5%) | 436(44.1%) | 738(46%) | 4664(46.6%) |
| Outdoors | 171(9.5%) | 139(9.9%) | 518(12.3%) | 97(9.8%) | 139(8.7%) | 1064(10.6%) |
| Total | 1796(100%) | 1407(100%) | 4220(100%) | 989(100%) | 1602(100%) | 10014(100%) |
| **Highest Education Level** |  |  |  |  |  |  |
| No education | 1071(59.6%) | 956 (68%) | 2734(64.8%) | 705 (71.3%) | 1164 (72.6%) | 6631 (66.2%) |
| Primary | 586(32.6%) | 378 (26.9%) | 1169 (27.7%) | 226 (22.9%) | 363 (22.7%) | 2722 (27.2%) |
| Secondary | 81 (4.5%) | 50(3.6%) | 223(5.3%) | 36(3.6%) | 49(3.1%) | 438(4.4%) |
| Higher | 58(3.2%) | 23(1.6%) | 94(2.4%) | 22(2.2%) | 26(1.7%) | 223(2.2%) |
| Total | 1796(100%) | 1407 (100%) | 4220 (100%) | 989 (100%) | 1602 (100%) | 10014(100%) |

| **Place of Residence** |  |  |  |  |  |  |
| --- | --- | --- | --- | --- | --- | --- |
| Urban | 228 (12.7%) | 127(9%) | 480(11.4%) | 83 (8.4%) | 130 (8.1%) | 1048(10.5%) |
| Rural | 1568(87.3%) | 1280(91%) | 3740(88.6%) | 905(91.6%) | 1472 (91.9%) | 8965.5(89.5%) |
| Total | 1796 (100%) | 1407 (100%) | 4220(100%) | 989(100%) | 1602(100%) | 10014(100%) |
| **Wealth index** |  |  |  |  |  |  |
| Poorest | 341(19%) | 337(23.9%) | 992.5(23.5%) | 297(30.1%) | 441(27.6%) | 2408.8(24.1%) |
| Poorer | 415.5 (23.1%) | 303(21.6%) | 950(22.5%) | 224.6 (22.7%) | 407.8(25.5%) | 2301(23%) |
| Middle | 344(19.1%) | 321 (22.8%) | 880(20.9%) | 215(21.8%) | 329 (20.6%) | 2089(20.9%) |
| Richer | 396(22%) | 250.5(17.8%) | 774(18.3%) | 148.5(15%) | 249(15.6%) | 1818(18.2%) |
| Richest | 300(16.7%) | 195(13.9%) | 623.6(14.8%) | 103(10.4%) | 174(10.9%) | 1396.6(13.9%) |
| Total | 1796 (100%) | 1407 (100%) | 4220 (100%) | 989(100%) | 1602(100%) | 10014(100%) |

The proportion of LBW among low polluting fuel users was 17% while the proportion of LBW among biomass fuel users was 26.2%. From the total children included in this study 2591 (25.9%) were LBW, and 9703(96.9%) infants belong to households using biomass fuels [Table 2]. Exposure to cooking smoke is greater when cooking takes place inside the house rather than in a separate building or outdoors. In Ethiopia, cooking is done in a separate building in 4664 (46.6%) of households. This figure is similar to the EDHS report which was 47%.

**Table 2.** Distribution of Birth size from mothers recall with fuel types and kitchen location.

| **Variable** | **Birth weight status** |  |  |
| --- | --- | --- | --- |
|  | <2500gram<br>(Low birth weight) | >2500 gram<br>(Not Low birth weight) | Total |
| **Fuel type** |  |  |  |
| Low polluting cooking fuels | 53(2.1%) | 258 (3.5%) | 311 (3.1%) |
| High polluting cooking fuels<br>(biomass fuels) | 2538(97.9%) | 7165(96.5%) | 9703(96.9%) |
| Total | 2591(100%) | 7423 (100%) | 10014 (100%) |
| **Kitchen location** |  |  |  |
| Inside the house | 1181(45.6) | 3105(41.8%) | 4286(42.8%) |
| In separate building | 1174(45.3%) | 3491 (47.0%) | 4665(46.6%) |
| Outdoors | 236 (9.1%) | 827 (11.1%) | 1063(10.6%) |
| Total | 2591 (100%) | 7423 (100%) | 10014 (100%) |

### Bivariate analysis

The Bivariate logistic regression analysis revealed that, there is a significant association between many potential predictor variables and infants birth size as indicated in [Table 3]. Infants born from mothers who live in households utilizing biomass fuels were 1.7 (OR, 95% CI 1.3, 2.3) times more likely to be born with low birth weight as compared to those using low polluting fuels. Newborn infants whose mother residing in households where food is cooked inside the house or in a separate building were found to be more likely to be LBW compared to infants born from mothers with a households where cooking is mostly done outdoors. Among other maternal factors, ages of the mother at first pregnancy, being moderate or mildly anemic, and lower BMI were among the risk factors for the child being LBW. Similarly, the sex of the child, children born in rural areas and those belonging to families with lower wealth index were at increased risk of being LBW [Table 3].

**Table 3.** Bivariate analysis of child size at birth with the type of fuel kitchen location and other variables.

| Variable | Low birth<br>weight | Normal birth<br>weight | Unadjusted Odds ratios (95%<br>CI) | p-value |
| --- | --- | --- | --- | --- |
| **Type of fuel** |  |  |  |  |
| High Pollution Fuels | 2538 | 7165 | 1.7 (1.3-2.3)* | <0.001 |
| Low Pollution Fuels | 53 | 258 | **1** |  |
| **Kitchen location** |  |  |  |  |
| Inside the house | 1181 | 3105 | 1.3 (1.1-1.6)* | <0.001 |
| In separate building | 1174 | 3490 | 1.2 (1.001-1.4)* | 0.045 |
| Outdoors | 236 | 828 | **1** |  |
| **Gender of the baby** |  |  |  |  |
| Male | 1162 | 4079 | 1 |  |
| Female | 1429 | 3344 | 1.5 (1.4-1.6)* | <0.001 |
| **Birth order number** |  |  |  |  |
| 1 | 500 | 1362 | **1** |  |
| 2 | 420 | 1239 | 0.9 (0.8-1.1) | 0.313 |
| 3 | 358 | 1074 | 0.90(0.8-1.1) | 0.235 |
| 4+ | 1313 | 3748 | 0.96 (0.8-1.1) | 0.463 |
| **Mothers age at birth** |  |  |  |  |
| <20 | 1739 | 4732 | **1** | **1** |
| 20-29 | 821 | 2599 | 0.86 (0.78-0.9)* | 0.002 |
| 30-49 | 31 | 92 | 0.9 (0.6-1.4) | 0.648 |
| **Maternal BMI** |  |  |  |  |
| Low | 528 | 1459 | 1.8 (1.2-2.8)* | 0.006 |
| Normal | 1931 | 5507 | 1.8(1.2-2.6)* | 0.008 |
| Overweight | 104 | 320 | 1.6 (1.01-2.6)* | 0.043 |
| Obese | 28 | 137 | **1** | **1** |
| **Maternal Anemia level** |  |  |  |  |
| Sevier | 46 | 101 | 1.4 (0.99-2.01) | 0.056 |
| Moderate | 202 | 499 | 1.2 (1.04-1.5)* | 0.014 |
| mild | 629 | 1553 | 1.2 (1.1-1.4)* | <0.001 |
| Not anemic | 1714 | 5270 | **1** | **1** |
| **Pregnancy intention when become pregnant** |  |  |  |  |
| Then | 1918 | 5633 | 0.9 (0.8-1.1) | 0.316 |
| Later | 461 | 1218 | 1.02 (0.8-1.2) | 0.843 |
| never | 212 | 572 | **1** | **1** |
| **Maternal Chat chewing** |  |  |  |  |
| No | 2137 | 6285 | **1** | **1** |
| Yes | 454 | 1138 | 1.2 (1.04-1.3)* | 0.009 |
| **Maternal Alcohol Drinking** |  |  |  |  |
| No | 1767 | 5334 | **1** | **1** |
| Yes | 824 | 2089 | 1.2 (1.1-1.3)* | <.001 |
| **Mothers education** |  |  |  |  |
| No education | 1869 | 4762 | 1.4 (1.03-2.0)* | 0.034 |
| Primary | 589 | 2133 | 1 (0.7-1.4) | 0.993 |
| Secondary | 85 | 353 | 0.9(0.6-1.3) | 0.484 |
| Higher | 48 | 175 | 1 | 1 |
| **Type of residence** |  |  |  |  |
| Urban | 214 | 835 | 1 | 1 |
| Rural | 2377 | 6588 | 1.4(1.2-1.6)* | <0.001 |
| **Wealth index** |  |  |  |  |
| Poorest | 739 | 1670 | 1.8 (1.5-2.1)** | <0.001 |
| Poorer | 632 | 1669 | 1.5 (1.3-1.8)** | <0.001 |
| Middle | 545 | 1545 | 1.4 (1.2-1.7)** | <0.001 |
| Richer | 398 | 1420 | 1.1 (0.95-1.3) | 0.188 |
| Richest | 277 | 1119 | 1 | 1 |

\* p < 0.05, \*\* p < 0.01, \*\*\* p < 0.001

### Multivariable analysis

We have analyzed the effect of fuel type and kitchen location on maternal report of birth size hierarchically. In the first model we have included and analyzed the effect of fuel type and kitchen location by including child factors. After that in the second model, we took the candidates from the first model were added with the maternal factors. In the final model we take all the candidates from the child and maternal factors and included with socio-demographic factors, one by one to the main exposure variables i.e. type of cooking fuel, and kitchen location. In the initial stage (model 1) when the impact of cooking fuel and kitchen location was assessed with child factors, on maternal report of birth weight, the use of biomass fuel had significant effect on maternal report of child size. Mothers who has used biomass fuels were 1.7 (adjusted OR 1.7; 95% CI 1.3, 2.3) times more likely to give birth to a LBW baby when compared to mother who has used low polluting fuels. Apart from the type of fuel type the kitchen location was also had a significant effect on maternal reported birth size. Cooking inside the house had (adjusted OR 1.3; 95% CI 1.15, 1.6) and cooking in a separate building had (adjusted OR 1.2; 95% CI 1.02, 1.4) effect on LBW respectively, when compared to households cooking outdoors. From the child factors being female gender was significantly associated with LBW other than fuel type and Kitchen location while birth order played no significant role on LBW [Table 4].

**Table 4.**
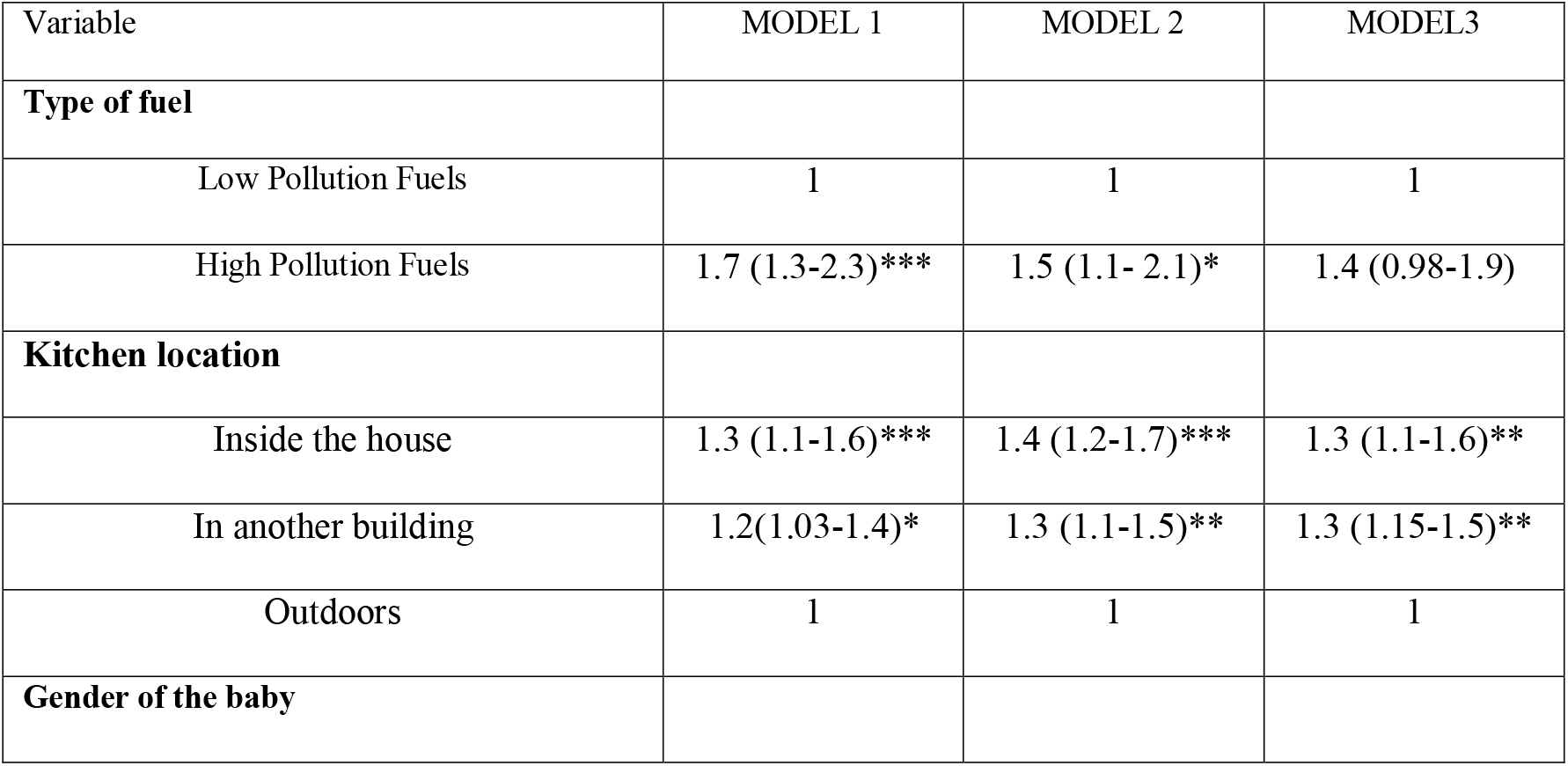
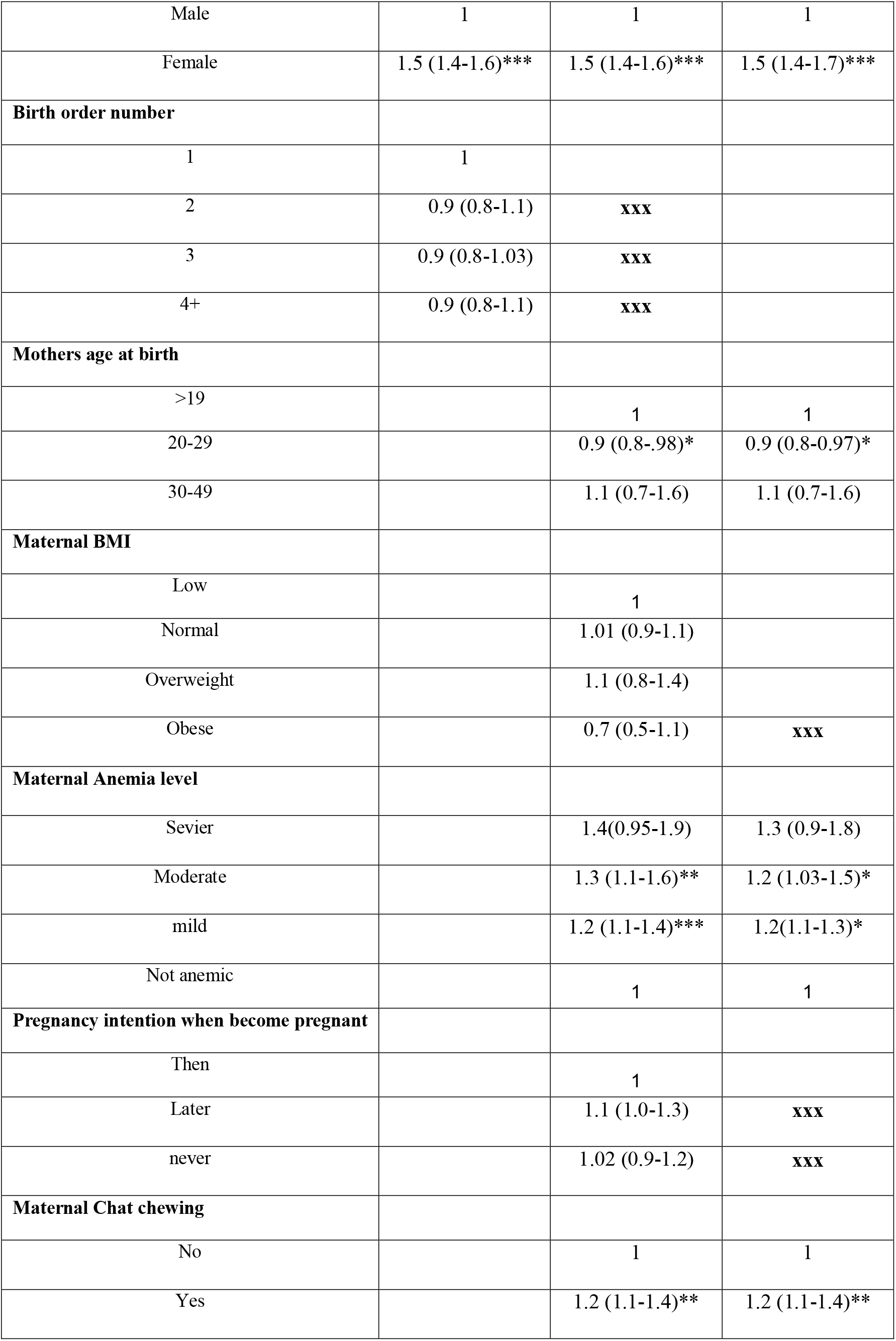
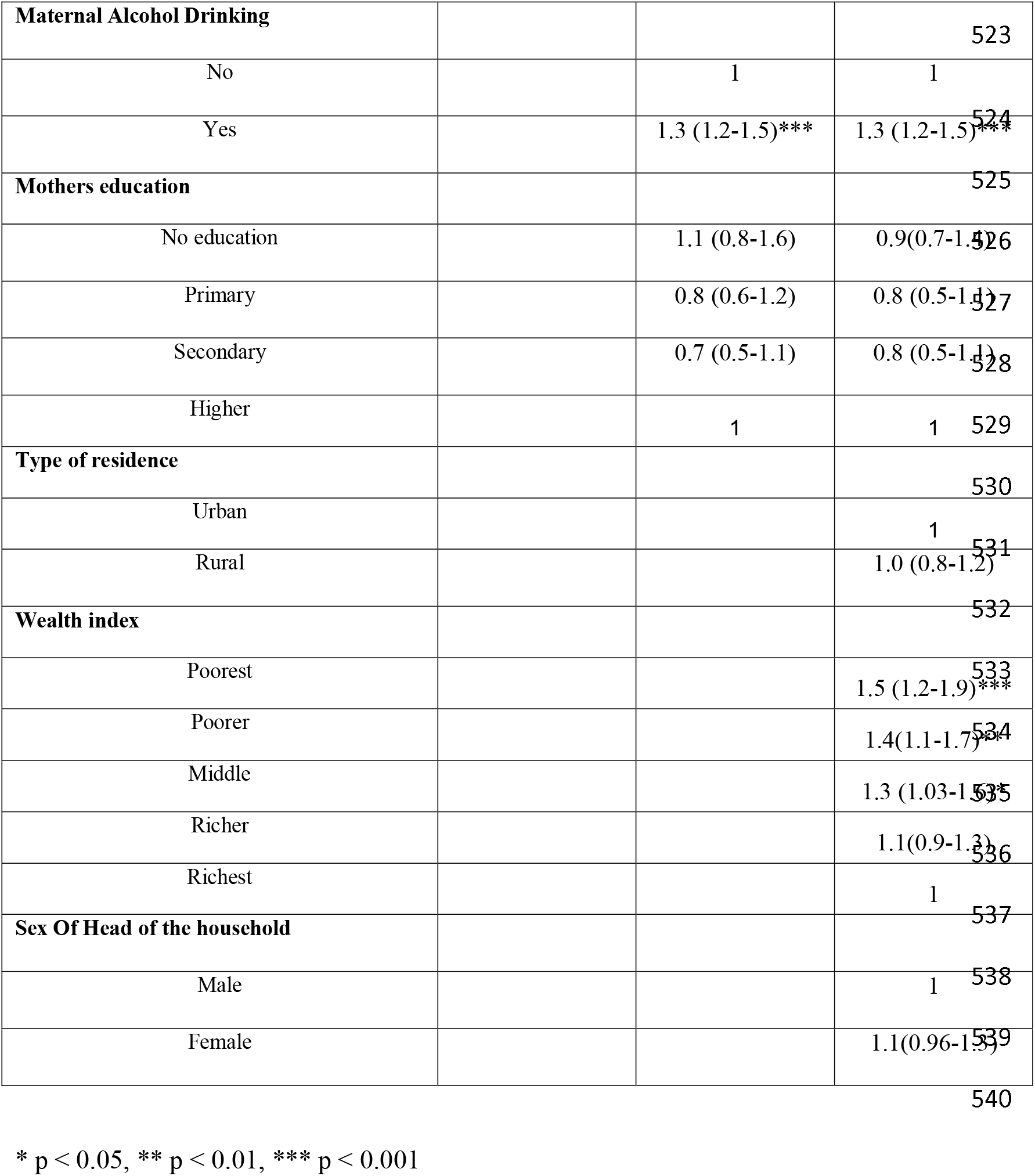
Multivariable analysis of child size at birth with fuel type, kitchen location and other variables.

In the second stage (model-2), the selected candidates from (model 1) were added to the maternal characteristics such as the age of the mother at first birth, maternal, educational status, BMI, Anemia level, maternal Chat chewing, and alcohol drinking. There was a reduction in the strength of the effect of fuel type on LBW but still the association was significant. Considering the child and maternal factors into consideration, biomass fuel use was significantly associated with LBW (adjusted OR 1.5; 95% CI 1.1, 2.1) while the strength of the effect of kitchen location on LBW showed a marginal increment at this stage. Other maternal factors significantly affecting the child’s birth size were maternal Anemia (*p* =<0.001), maternal chat chewing (*p*=0.002), and maternal Alcohol drinking (*p*<0.001). Being a female baby still had a significant effect on LBW in Model 2 and its effect size was unchanged at this stage. In final model (model-3) child, maternal, and demographic variables were included together with the exposure variables to examine the effect on LBW. At this stage, mothers who live in households using biomass fuels tend to have a higher odds of giving a LBW child but the association was insignificant (*p* = 0.069) (adjusted OR 1.4, 95% 0.98, 1.9), but the kitchen location where the food is prepared for the household remains a significant predictor of LBW. Cooking either in the household (p=0.001) (Adjusted OR, 1.3; 95% CI 1.1, 1.6) or in a separate building (*p*=0.002) (Adjusted OR; 95% CI 1.3 1.2, 1.5) were, almost equally found to have a significant effect on birth size compared to cooking food outside the household. Being a female child was not only a significant predictor in the final model; its strength was found somewhat similar and strong in all the models. Maternal age, Anemia level, pregnancy intention, chat chewing, alcohol drinking, and wealth index were also found to be significant predictors of the child’s birth size in the final model (Tab. 3). However, the types of residence, sex of head of the household and maternal education were not associated with birth weight in the final model. The wealth index might have masked the association between cooking fuel and birth weight in the final model. This may imply that people with higher incomes could afford to buy cleaner fuels which are usually more expensive.

## Discussion

According to the descriptive findings of the study, a quarter of children were born with low birth weight and about 62% of children with low birth weight were very small and 38% smaller than average. This finding is comparable to finding from rural India of National Family Health Survey [38] which found the level of low birth weight to be 23% and another finding from India which identified 23.8% in Dehradun [39], and 23% in Rural Karnataka [40].

On the other hand, only three percent of the households use low polluting fuel and the main source of fuel item for the majority of the households (about 85%) is wood. This level of using highly polluting cooking fuels among households in Ethiopia is much higher than the finding from India 72.9% [28], Malawi, 80% [30], and Ghana [41] which have found that 66% of households using biomass fuel as their main cooking fuel.

Regarding the association between birth weight and fuel type, children of mothers from households with high polluting fuels were about two times more likely to have low birth weight on bivariate analysis and intermediate models. However, the use of biomass fuel was not significantly associated with birth weight after all the confounding variables were controlled on multivariate analysis of the final model. A similar finding was revealed from a study by WHO on indoor air pollution from biomass fuel use and risk of LBW [42] and study from Ghana [41]. Another study from Malawi using DHS data has revealed an increased but insignificant association between biomass fuel use and low birth weight [30]. Similarly, a study conducted using Indian DHS data, identified a higher risk of low birth weight from biomass fuel use at bivariate analysis but insignificant relationship after adjustment for other predictor variables [28]. However, other findings from different studies showed that using biomass fuels was associated with a higher risk of low birth weight. In this regard, using biomass fuel was also implicated with twofold increased risk on low birth weight in Lanzhou, China [43] and 175g reduction in mean birth weight was observed in children born to mothers that used biomass fuels in Zimbabwe. A similar finding was reported using the Pakistan Demographic and Health Survey data, where children born from households with biomass fuel (wood) users were found 41% more likely to have LBW compared to children born from households using cleaner fuel types such as natural gas [29]. In the current study, the association between birth weight and fuel type might be masked by the effect of the wealth index of the households since the richer households might have the capability to afford those clean fuel types which are more expensive in the Ethiopian context. The strength of the association wealth index has on birth size was decreasing as we go from the poorest to the richest which could be a clear indication that most solid biomass fuels are either cheap or free (in the case of agricultural residues and animal dungs) compared to low polluting fuel types such as electricity, liquid petroleum gas, natural gas and biogas which are relatively expensive.

This study identified that, cooking whether inside the house or in a separate building was associated with the decreased maternal report of child size, compared to cooking outdoors. The finding of the practice of cooking food in the house with LBW was consistent with a study conducted in the Wolaita zone, southern Ethiopia [35] and Dang district of Nepal [44], where Children born to mothers regularly cooking inside the house were found to report a low birth weight baby as compared to those mothers cooking outdoor. This might be due to risks associated with the type of fuel use, exposure time, ventilation status and the efficiency of the cooking stove. The world health organization set a public health standard for indoor air pollutants (24-hour mean: PM_2.5_ < 25 μg/m^3^ and PM_10_ <50 μg/m^3^) which can be attained through the use of cleaner fuels, well ventilated households and kitchens, efficient cookstoves and less exposure times. Since none or most of them are absent in most developing countries including Ethiopia, biomass fuel use together with longer exposure times, poor ventilation and unimproved cooking stoves might be path ways for maternal exposure to higher level of pollutants which in turn results in LBW [15, 16, 23-24].

According to our findings cooking inside the house and cooking in a separate building has an almost similar effect on birth size when compared to cooking outdoors. This is a clear indication that, unless efficient stoves are in place with adequate ventilation when using solid biomass fuels the effect of cooking inside the house or in a separate kitchen might not have a difference. This is apparent in countries such as Ethiopia, where widespread use of improved cookstove has been deterred by, different factors such as the meager income of the poor to afford, lack of infrastructure, sluggish market penetration into remote villages, lack of know-how of utilization, and information gap [45]. Our finding was in contrary to similar researches conducted in Bangladesh [27], where the cooking place was not significantly associated with child size at birth. One reason for the difference might be due to the difference in the location, ventilation status and other kitchen characteristics. It could be also attributed to the proportion of efficient stoves users, proportion of solid biomass fuel users and other socio-economic characteristics of the study population.

According to this study finding, wealth index was found to be a significant predictor of LBW. Newborns from poorer households were found to be at higher risk of being low birth weight than newborns from richer households. Ethiopian Female neonates were also lighter than their male counterparts. This finding was similar to findings from other studies elsewhere including studies from Japan [46], Northern Ethiopia [33]. On the other, hand neonates of anemic women had a marginally higher possibility of being LBW compared to those of non-anemic women. From a study conducted in Northern Ethiopia, anemic women were found to be nine times more likely to deliver an infant with low birth weight [33] and similar finding was found from a study of national Family Health Survey-IV in India [28]. Similarly, marginal association was found between maternal chat chewing and birth weight. This finding also complies with finding from study conducted in Yemen [47].

### Methodological Strengths and Limitations

The strength of this study is that the findings can be generalized at the country level since the study utilized data from a nationally representative household survey. However, children were classified as LBW or not LBW based on the mothers subjective judgment which might introduce measurement bias on the outcome ‘low birth weight’. This could have implications on the result. Apart from the reliance on self-report of birth size, the cross-sectional design might pose problems in establishing the temporal link between exposure and outcome. This study assumed that maternal exposure to biomass fuels was a phenomenon occurred repeatedly for a long time before pregnancy, which might not always be the case. Besides, households may not use the same fuel types all the time. Changes in use from biomass fuel use to cleaner fuel types might occur in real life situation that may lead to misclassification of the use of fuel types which in turn may compromise exposure and end up in under or overestimated effect. Therefore, the exclusion of the use of mixed fuel types and its effect on LBW is the biggest limitation of this study and the interpretation of the result must be with great caution.

## Conclusion

Use of biomass fuel and kitchen location was associated with child size at birth. Our finding has important program and policy implications for countries such as Ethiopia, where large proportions of the population rely on polluting biomass fuels for cooking. Furthermore, future studies should investigate the association using more direct methods for measurement of exposure to smoke emitted from biomass fuels on birth weight.

## Data Availability

The data are available from the Demographic and Health Survey program website. This data is publicly available online and it can be accessed at the following website by selecting the specific country Ethiopia. http:// dhsprogram.com/data/available-datasets.cfm

## Acknowledgment

Authors are grateful to DHS ICF, USA for giving us permission to use the EDHS 2016 dataset.

## Data Availability Statement

The data are available from the Demographic and Health Survey program website. This data is publicly available online and it can be accessed at the following website by selecting the specific country Ethiopia. http://dhsprogram.com/data/available-datasets.cfm

## Authors’ contributions

**Conceptualization:** Girum Gebremeskel Kanno **Formal analysis:** Girum Gebremeskel Kanno and Sewitemariam Desalegn Andarge **Methodology:** Girum Gebremeskel Kanno and Sewitemariam Desalegn Andarge **Project administration:** Girum Gebremeskel Kanno **Supervision:** Adane Tesfaye Anbesse, Mohamed Feyisso Shaka and Miheret Tesfu Legesse **Writing – original draft:** Girum Gebremeskel Kanno and Mohamed Feyisso Shaka **Writing – review & editing:** Girum Gebremeskel Kanno, Mohamed Feyisso Shaka and Adane Tesfaye Anbesse, Miheret Tesfu Legesse

## References

1. World Health Organization: Global Nutrition Targets 2025: Low birth weight policy brief. 2014.

2. Christian P., Lee S. E., Angel M. D., Adair, L. S., Arifeen S. E, Ashorn P., et al. Risk of childhood undernutrition related to small-for-gestational age and preterm birth in low- and middle-income countries. Int J Epidemiol. 2013; 42(13): 40–55.

3. World Health Organization: Comprehensive implementation plan on maternal, infant and young child nutrition. Resolution WHA65.6. 2012.http://www.who.int/nutrition/topics/WHA65.6_resolution_en.pdf?ua=1, accessed 17

4. Ethiopian Central Statistical Agency: Ethiopian Demographic and Health Survey report: Key Indicators Report. The DHS Program ICF. 2016.

5. Kim D. and Saada A. The Social Determinants of Infant Mortality and Birth Outcomes in Western Developed Nations: A Cross-Country Systematic Review. Int. J. Environ. Res. Public Health 2013; 10: 2296-2335; doi:10.3390/ijerph10062296

6. Gu H., Wang L., Liu L., Luo X., Wang J., Hou F., Denis P. et al.. A gradient relationship between low birth weight and IQ: A meta-analysis. Sci Rep. 2017; 7(1):18035. doi:10.1038/s41598-017-18234-9

7. Jornayvaz F. R., Vollenweider P., Bochud M., Mooser V., Waeber G. and Marques-Vidal P. Low birth weight leads to obesity, diabetes and increased leptin levels in adults: the CoLaus study. Cardiovasc Diabetol. 2016; 15: 73. DOI 10.1186/s12933-016-0389-2

8. Risnes K. R., Vatten L. J, Baker J. L, Jameson K., Sovio U., Kajantie E., et al. Birthweight and mortality in adulthood: a systematic review and meta-analysis. Int J Epidemiol. 2011; 40(3):647–61. Doi: 10.1093/ije/dyq267.

9. Larroque B, Bertrais S., Czernichow P., Léger J. School difficulties in 20-year-olds who were born small for gestational age at term in a regional cohort study. Pediatrics. 2001;108(1):111–115. Doi: 10.1542/peds.108.1.111

10. UNICEF-WHO: Low Birthweight Estimates Levels and trends 2000–2015. 2019.

11. Boy E., Bruce N., and Delgado H. Birth Weight and Exposure to Kitchen Wood Smoke during Pregnancy in Rural Guatemala. Environ. Health Perspect. 2002;110:1.

12. Mishra V., Dai X., Smith K. R., and Mika L. Maternal Exposure to Biomass Smoke and Reduced Birth Weight in Zimbabwe. Ann. Epidemiol. 2004; 14(10):740–747 doi:10.1016/j.annepidem.2004.01.009

13. Kim KH, Jahan SA & Kabir E A review of diseases associated with household air pollution due to the use of biomass fuels. J. Hazard. Mater. 2011; 192: 425–431.

14. Balakrishnan K. Sambandam S. Ramaswamy P. Mehta S. Smith KR. Exposure assessment for respirable particulates associated with household fuel use in rural districts of Andhra Pradesh, India. J Expo Anal Environ Epidemiol. 2004;14: 14-25. DOI: 10.1289/ehp.9479

15. Smith KR. Indoor air pollution in developing countries: recommendations for research. Indoor Air. 2002; 12: 198–207.

16. Oliveira BF, Ignotti E, Hacon SS. A systematic review of the physical and chemical characteristics of pollutants from biomass burning and combustion of fossil fuels and health effects in Brazil. Cad Saude Publica. 2011; 27(9):1678-98. DOI: http://dx.doi.org/10.1590/S0102-311X2011000900003version=html

17. Zhang J.J. and Smith K.R. Household Air Pollution from Coal and Biomass Fuels in China: Measurements, Health Impacts, and Interventions. Environ. Health Perspect. 2007;115(6):848–55. Doi: 10.1289/ehp.9479

18. Ha E.H., Hong Y.C., Lee B.E., Woo B.H., Schwartz J. and Christiani D. C. Is Air Pollution a Risk Factor for Low Birth Weight in Seoul? Epidemiology. 2001; 12(6):643-648. DOI: 10.1097/00001648-200111000-00011

19. Maisonet M., Bush T. J., Correa A. and Jaakkola J.K. Relation between Ambient Air Pollution and Low Birth Weight in the Northeastern United States. Environ. Health Perspect. 2001;109(3):351–356.

20. Washam C. Cooking with wood may fuel low birth weight: kitchen smoke puts babies at risk. Environmental health perspectives. 2008; 116(4):A173–A.

21. Ritz B, Yu F. The effect of ambient carbon monoxide on low birth weight among children born in southern California between 1989 and 1993 Environ. Health Perspect. 1999; 107(1):17–25. Doi: 10.1289/ehp.9910717

22. World Health Organization; Opportunities for transition to clean household energy. application of the Household Energy Assessment Rapid Tool (HEART) in Ethiopia. Getachew E Beyene, Abera Kumie, Rufus Edwards, Karin Troncoso. 2018; ISBN 978-92-4-151449-1

23. Bonjour S., Rohani H. A., Wolf J., Bruce N. G, Mehta S., Prüss-Ustün A., Lahiff M., Rehfuess E. A., Mishra V., Smith K. R. Solid fuel use for household cooking: country and regional estimates for 1980-2010. Environ Health Perspect. 2013;121(7):784-90. DOI: 10.1289/ehp.1205987

24. Amegah AK, Quansah R, Jaakkola JJK. Household Air Pollution from Solid Fuel Use and Risk of Adverse Pregnancy Outcomes: A Systematic Review and Meta-Analysis of the Empirical Evidence. PLoS ONE. 2014; 9(12): e113920. https://doi.org/10.1371/journal.pone.0113920

25. UNICEF; Levels and Trends in Child Mortality: Estimates Developed by the UN Inter-Agency Group for Child Mortality Estimation (IGME). UNICEF: New York, 2015.

26. Lawn JE, Blencowe H, Oza S, You D, Lee A CC, Waiswa P et al. Every Newborn: progress, priorities, and potential beyond survival. Lancet. 2014; 384 (9988): 189–205.

27. Haider MR, Rahman MM, Islam F, Khan MM Association of Low Birthweight and Indoor Air Pollution: Biomass Fuel Use in Bangladesh. JH&P. 2016;6(11): 18–25. https://doi.org/10.5696/2156-9614-6-11.18

28. Sreeramareddy CT, Shidhaye RR and Sathiakumar N. Association between biomass fuel use and maternal report of child size at birth - an analysis of 2005-06 India Demographic Health Survey data. BMC Public Health. 2011; 11:403http://www.biomedcentral.com/1471-2458/11/403

29. Ahmed Z, Zafar M, Khan NA, Qureshi MS. Exposure to biomass fuel and low child birth weight – Findings of Pakistan Demographic and Health Survey 2006–2007. Int J Health Syst Disaster Manage. 2015;3:S19–26.

30. Milanzi EB, Namacha NM. Maternal biomass smoke exposure and birthweight in Malawi: Analysis of data from the 2010 Malawi Demographic and Health Survey. Malawi Med J. 2017;29(2):160–165.

31. Bekela MB, Shimbre MS, Gebabo TF, Geta MB, Tonga AT, Zeleke EA, Sidemo NB, and Getnet AB Determinants of Low Birth Weight among Newborns Delivered at Public Hospitals in Sidama Zone, South Ethiopia: Unmatched Case-Control Study. Hindawi Journal of Pregnancy. 2020; Article ID 4675701, https://doi.org/10.1155/2020/4675701

32. Talie A, Taddele M, and Alemayehu M. Magnitude of Low Birth Weight and Associated Factors among Newborns Delivered in Dangla Primary Hospital, Amhara Regional State, North west Ethiopia. Hindawi Journal of Pregnancy. 2017; Article ID 3587239, 6 pages https://doi.org/10.1155/2019/3587239

33. Asmare G, Berhan N, Berhanu M and Alebel A. Determinants of low birth weight among neonates born in Amhara Regional State Referral Hospitals of Ethiopia: unmatched case control study. BMC Res Notes. 2018; 11:447 https://doi.org/10.1186/s13104-018-3568-2

34. Sema A, Tesfaye F, Belay Y, Amsalu B, Bekele D, and Desalew A. Associated Factors with Low Birth Weight in Dire Dawa City, Eastern Ethiopia: A Cross-Sectional Study. Hindawi BioMed Research International. 2019; Article ID 2965094, https://doi.org/10.1155/2019/2965094

35. Admasie A, Kumie A and Worku A. Association of Household Fuel Type, Kitchen Characteristics and House Structure with Child Size at Birth in Wolaita Sodo, Southern Ethiopia. Open Publ Health J. 2018; 11: 298–308.

36. Wachamo TM, Bililign YN, Bizuneh AD Risk factors for low birth weight in hospitals of North Wello zone, Ethiopia: A casecontrol study. PLoS ONE. 2019; 14(3): e0213054. https://doi.org/10.1371/journal.pone.0213054

37. Alemu T, Umeta M. Prevalence and Predictors of “Small Size” Babies in Ethiopia: In-depth Analysis of the Ethiopian Demographic and Health Survey 2011. Ethiop J Health Sci. 2016; 26(3): 243–250.

38. Noor N, Kural M, Joshi T, Pandit D, Patil A. Study of maternal determinants influencing birth weight of newborn. Arch Med Heal Sci 2015;3:239. https://doi.org/10.4103/2321-4848.171912.

39. Negi KS, Kandpal SD, Kukreti M. Epidemiological factors affecting low birth weight. JK Sci 2006;8:31–4.

40. Metgud CS, Naik VA, Mallapur MD. Factors affecting birth weight of a newborn - a community based study in rural Karnataka, India. PLoS One 2012;7. https://doi.org/10.1371/journal.pone.0040040.

41. Weber E, Adu-bonsaffoh K, Vermeulen R, Klipstein-grobusch K, Grobbee DE, Browne JL, et al. Household fuel use and adverse pregnancy outcomes in a Ghanaian cohort study. Reprod Health 2020;17:1–8.

42. World Health Organization. Indoor air pollution from solid fuels and risk of low birth weight and stillbirth. Annu. Conf. Int. Soc. Environ. Epidemiol., Johannesburg: 2007, p. 1–39.

43. Jiang M, Qiu J, Zhou M, He X, Cui H, Lerro C, et al. Exposure to cooking fuels and birth weight in Lanzhou, China: A birth cohort study. BMC Public Health 2015;15:1–10. https://doi.org/10.1186/s12889-015-2038-1.

44. K. C. A, Basel PL, Singh S (2020) Low birth weight and its associated risk factors: Health facility-based case-control study. PLoS ONE 15(6): e0234907. https://doi.org/10.1371/journal.

45. Dawit Diriba Guta Assessment of Biomass Fuel Resource Potential And Utilization in Ethiopia: Sourcing Strategies for Renewable Energies. Int. J. Renew. Energy Res. 2012; 2(1).

46. Terada M, Matsuda Y, Ogawa M, Matsui H, Satoh S. Effects of maternal factors on birth weight in Japan. J Pregnancy 2013;2013. https://doi.org/10.1155/2013/172395.

47. Abdel-Aleem, A.M, Abdulkader A.A, Mustafa M, S.A, Nasr A A, Assad, A M.M. Khat Chewing During Pregnancy: an Insight on an Ancient Problem. Impact of Chewing Khat on Maternal and Fetal Outcome among Yemeni Pregnant Women. J Gynecol Neonatal Biol. 2015; 1(2). DOI: 10.15436/2380-5595.15.004

